# Health-Related Quality of Life and psychological distress of adolescents during the COVID-19 pandemic in Geneva

**DOI:** 10.1101/2021.09.20.21263812

**Authors:** Roxane Dumont, Viviane Richard, Hélène Baysson, Elsa Lorthe, Giovanni Piumatti, Stephanie Schrempft, Ania Wisniak, Rémy P. Barbe, Klara M. Posfay-Barbe, Idris Guessous, Silvia Stringhini, on behalf of the Specchio-COVID19 study group

## Abstract

**Purpose:** Our objective was to assess adolescent’s Health-Related Quality of Life (HRQoL) and psychological distress, from their own and their parents’ perspective, and to examine associated risk factors during the COVID-19 pandemic in Geneva, Switzerland.

**Methods:** A random sample of adolescents, aged 14-17 years, and their families was invited to a serosurvey in November and December 2020. Adolescents’ HRQoL was evaluated using the validated adolescent-reported KIDSCREEN-10 and parent-reported KINDL^®^ scales. Psychological distress was assessed with self-reported sadness and loneliness, and using the KINDL^®^ emotional well-being scale. Risk factors for adolescents’ low HRQoL and psychological distress were identified using generalized estimating equations and both adolescents’ and their parents’ perceptions were compared.

**Results:** Among 240 adolescents, 11% had a low HRQoL, 35% reported sadness and 23% reported loneliness. Based on parents’ perception, 12% of the adolescents had a low HRQoL and 16% a low emotional well-being. Being a girl (aOR=3.29; 95%CI: 1.64-6.57), increased time on social media (aOR=2.05; 95%CI: 1.08-3.88), parents’ *average to poor* mood (aOR=2.81; 95%CI: 1.21-6.56) and *average to poor* household financial situation (aOR=2.30; 95%CI: 1.00-5.29) were associated with an increased risk of sadness. Mismatches between adolescents’ and their parents’ perception of HRQoL were more likely for girls (aOR**=**2.88; 95%CI: 1.54-5.41) and in households with lower family well-being (aOR**=**0.91; 95%CI: 0.86-0.96).

**Conclusion:** A meaningful proportion of adolescents experienced low well-being during the second wave of COVID-19. Adolescents living in underprivileged or distressed families seemed particularly affected. Monitoring is necessary to evaluate the long-term effects of the pandemic on adolescents.

**Implications and Contribution:** This study describes the psychological well-being of a population-based sample of adolescents in Geneva, Switzerland amid the COVID-19 pandemic, and identifies adolescents at risk of distress. This study provides further insight by comparing adolescents’ well-being as reported by themselves and their parents.

## 1. INTRODUCTION

The COVID-19 pandemic and the measures put in place by public health authorities to contain its spread have caused significant disruptions in daily life and raised concerns for mental health of the entire population, and in particular adolescents [1]. Adolescence, defined as the developmental phase from childhood to adulthood [1,2], is characterized by important psychological and physical changes, greater vulnerability to external events, and less resources to adapt to hard circumstances [3]. During this sensitive stage of life, the importance of peer-interactions increases in parallel with a desire for greater autonomy from parents [4].

Because of increased difficulties to meet these developmental needs, adolescents may have been particularly affected by the pandemic [5], despite being at low risk of severe SARS-CoV-2 infection [6]. Educational disruptions due to school closures or quarantine, the widespread use of distance learning, the introduction of social distancing, or cancellation of extra-curricular activities combined with the general stay-at-home message, led to significant changes in daily life, less time spent with peers, and more with the family. This situation could have exacerbated tensions within the family or, represented an opportunity to strengthen family relationships [7]. These unprecedented circumstances also resulted in changes in health behaviours among adolescents such as an increase in screen time [8] and “junk food” consumption [9], together with a decrease in physical activity [10]. Adolescents may have been overly burdened by these life changes along with the worrying pandemic environment, as confirmed by a narrative review showing that adolescents’ mental health was negatively impacted by the COVID-19 pandemic situation [11]. Adolescents living in underprivileged families or whose parents present a decreased mental or health state, including a SARS-CoV-2 infection, could be particularly vulnerable as they may have less support and resources to adapt to the changes induced by the pandemic [12,13].

Sanitary restrictions and mortality due to the COVID-19 pandemic were relatively light in Switzerland compared with other countries [14], which may have resulted in a reduced impact on adolescents. After a first 10 week lockdown in spring 2020, schools remained open, while restrictions such as limitations on size of gatherings, remote work and mask wearing in public spaces from 12 years old were maintained. Inland travel was never limited; non-essential shops, sport facilities and cultural places intermittently closed according to the epidemiological situation. Yet, it is important to assess adolescents’ well-being and psychological distress in this specific context to be able to internationally compare the impact of different measures. A better understanding of adolescents’ well-being during this period is also useful for the design of new measures as the COVID-19 pandemic continues.

In this study, we assess adolescents’ health related quality of life (HRQoL) and psychological distress, as well as their risk factors. We also compare their own and their parents’ perceptions. Data comes from a population-based sample of adolescents in the canton of Geneva, Switzerland, primarily recruited for a SARS-CoV-2 serosurvey during the second COVID-19 wave.

## 2. METHODS

### Survey design

A serological study was conducted between November 23rd and December 23rd 2020, in the general population of the canton of Geneva, Switzerland [15]. Children and adolescents aged 6 months to 17 years were randomly selected from the Swiss Federal Statistical Office’s registers and were invited with their family members for a free anti-SARS-CoV-2 serology. 597 families participated (participation rate: 17%). Of these, 194 families had adolescents aged between 14 and 17 years, and were included in the current study. This age range was chosen for adolescents to be mature enough to autonomously answer the study questionnaire.

Each family designated a “referent parent” who completed comprehensive health and socio-demographic online questionnaires about themselves and their child(ren). Additionally, adolescents were asked to complete a paper questionnaire about their well-being and life habits since the start of the COVID-19 pandemic, mid-March 2020. This questionnaire was completed during the serology appointment in a separate area ensuring confidentiality, especially from parents. Adolescents were included in this analysis if the three above-mentioned questionnaires were completed (Figure 1).

**Figure 1:**
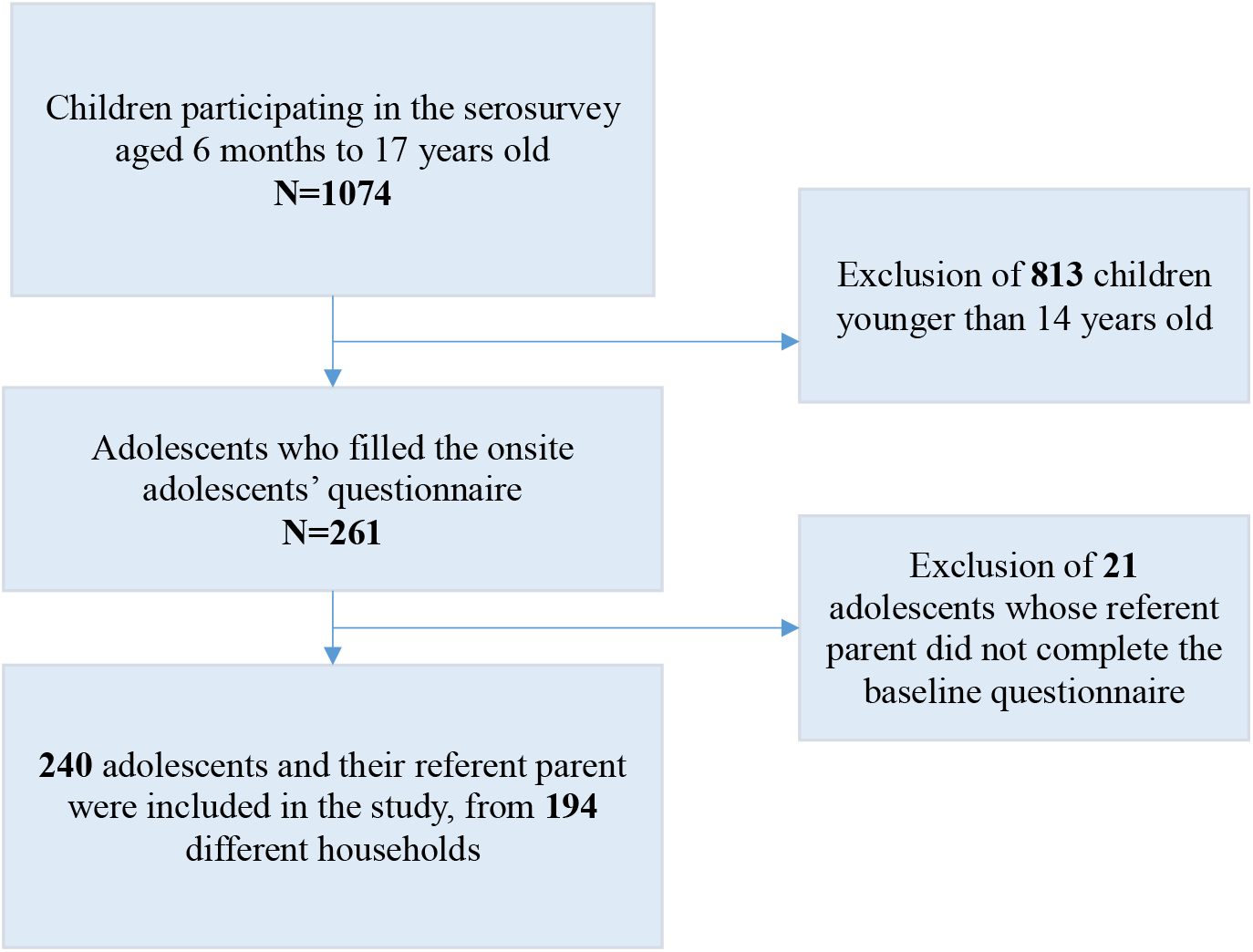
Study flow diagram

### Measures

#### HRQoL and psychological distress - adolescent’s perception

HRQoL as perceived by the adolescents was evaluated over the previous week using the validated French version of the standardized KIDSCREEN-10 scale [16]. KIDSCREEN-10 evaluates HRQoL in children and adolescents aged 8 to 18 years based on 10 items, enabling a normalized score calculation from 0 to 100. The internal consistency of the overall self-reported KIDSCREEN-10 index was good (α=0.81). Adolescents with a HRQoL score lower than one standard deviation below the study population mean were considered as having a low HRQoL [17]. The 2 items of the KIDSCREEN-10 targeting mood and feelings (sadness and loneliness) were separately used as binary proxies for psychological distress. Adolescents were considered to feel sad or lonely if they reported feeling so “quite often”, “very often” or “always” over the last week.

#### HRQoL and psychological distress - parent’s perception

The parent’s perception of their children’s well-being over the previous week was based on the French version of the KINDL^®^ for parents [18]. This scale measures HRQoL of children and adolescents aged from 7 to 17 years based on their parent’s answers, by combining 24 items covering 6 dimensions: physical well-being, emotional well-being, self-esteem, family, friends and school. Adolescents with an overall HRQoL score lower than one standard deviation below the study population mean were considered as having a low HRQoL [17]. Overall internal consistency of the KINDL^®^ was high (α=0.85). Focusing on the distinct KINDL^®^ dimensions, psychological distress was defined as an emotional well-being score lower than one standard deviation below the study population mean. This dimension showed acceptable consistency (α=0.69).

#### Outcomes

The 5 outcomes of interest were HRQoL from both the adolescents’ and the parents’ perception, as well as self-reported sadness and loneliness, and emotional well-being reported by the referent parent.

#### Potential risk factors

Potential risk factors were identified based on a literature review and descriptive analyses. Age, sex, screen time (hours per day), change in time spent on social networks since the start of the pandemic (increase, decrease, no change) and anti-SARS-CoV-2 serological status of the adolescent were selected, as well as age, sex, education, self-perceived mood and anti-SARS-CoV-2 serological status of the referent parent. Values of the anti-SARS-CoV-2 serology ≥0.8 µ/mL were considered positive (Elecsys anti-SARS-CoV-2 S; Roche Diagnostics, Rotkreuz, Switzerland [19]); the serological assessment was conducted before the start of the vaccination campaign in Switzerland. Parent education was measured with a three-level scale: lower (compulsory or no formal education), middle (secondary education), and higher (tertiary education). Self-perceived mood of the parent was defined as good if the answer to the question “In general, outside the pandemic context, how would you assess your mood ?” was “good” or “very good”, and average to poor for answers such as “average”, “poor” or “very poor”. Household level variables such as household size, density, financial situation and family well-being were also included. Household size was expressed as the number of people living in the household. Household density was defined using the household crowded index [20], calculated as the household size divided by the number of bedrooms. Households with a ratio higher than 1.5 were considered crowded. Household financial situation was considered as good if the referent parent answered that they could save money or face minor unexpected expenses, and average to poor if they selected one of the following statements: “I have to be careful with my expenses and an unexpected event could put me into financial difficulty” or “I cannot cover my needs with my income and I need external support”. Family well-being was measured using the family dimension of the KINDL^®^ scale for parents (α=0.71), which focuses on the relationship between the children and their parents [18].

### Statistical analysis

After excluding participants with missing data, multivariable models were performed for each outcome with adolescents’ age, sex, anti-SARS-CoV-2 serological status, screen time and change in time spent on social network, referent parents’ age, sex, anti-SARS-CoV-2 serological status and mood, as well as household size, density and financial situation as predictors. As some of the adolescents were siblings, a generalized estimating equation (GEE) function [21] was used to correct for the familial dependency in the observations with a covariance matrix defined as “exchangeable” and tests based on sandwich-corrected robust standard errors. Multivariable model results were reported as adjusted odds ratios (aOR) with 95% confidence intervals (95%CI).

Discrepancies between self-reported and parent-reported low HRQoL were coded as a binary variable: 1 if perceptions were different and 0 if they were similar. Risk factors of discrepancy were assessed with GEE adjusting for the above-mentioned covariates, as well as the family well-being score from the KINDL^®^ scale.

Statistical significance was defined at a level of confidence of 95% and all analyses were performed with R (version 4.0.3).

## 3. RESULTS

The sample consisted of 240 adolescents from 194 households. Mean age was 15 years (SD 2.1 years) and 47% were females. Referent parents’ mean age was 47 years (SD 9.0 years), 75% being mothers. Table 1 presents a descriptive overview of the adolescents’ and their parents’ socio-demographic characteristics.

**Table 1:**
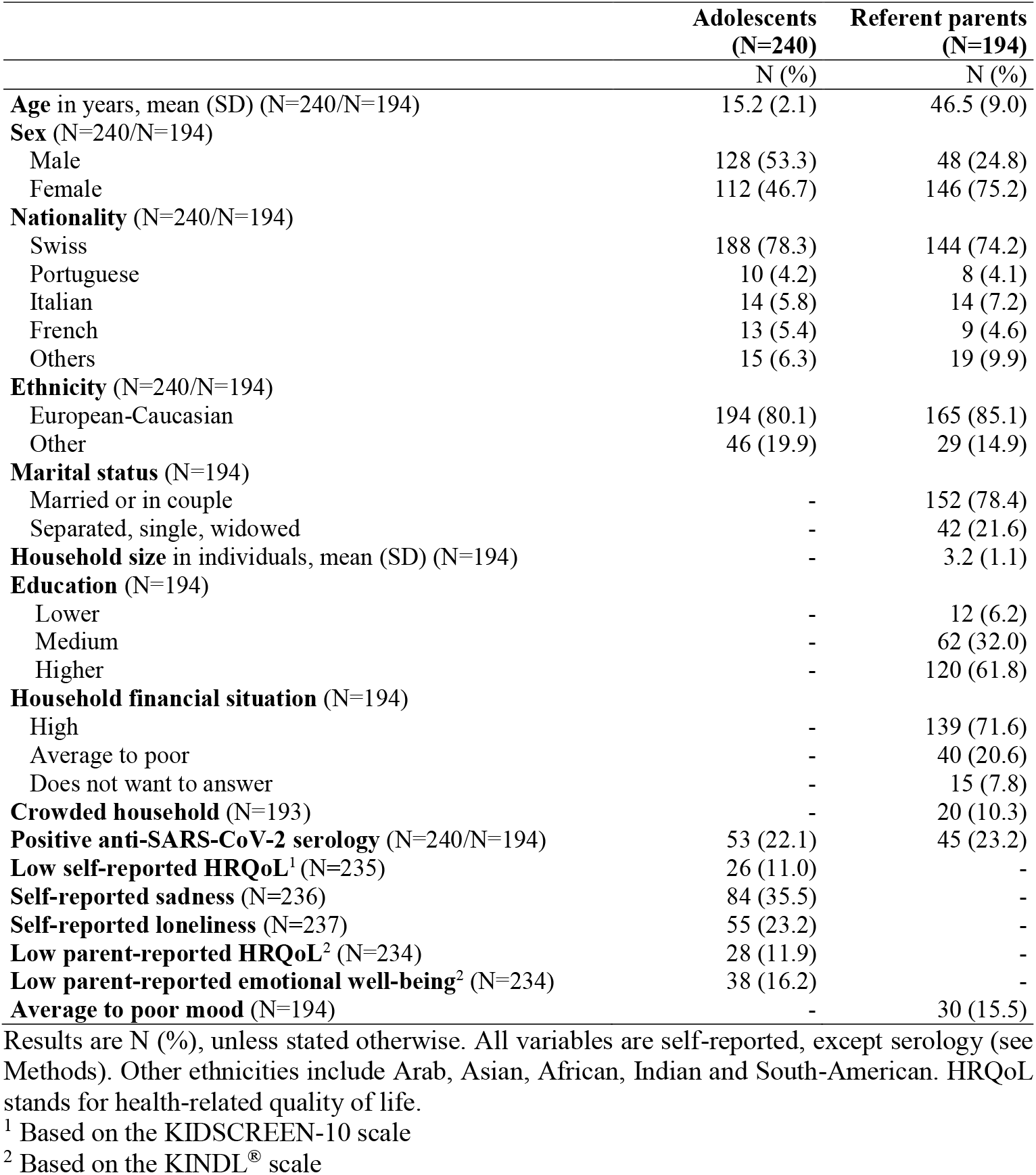
Characteristics of the study population

|  | **Adolescents**<br>**(N=240)** | **Referent parents**<br>**(N=194)** |
| --- | --- | --- |
|  | **N (%)** | **N (%)** |
| **Age** in years, mean (SD) (N=240/N=194) | 15.2 (2.1) | 46.5 (9.0) |
| **Sex** (N=240/N=194) |  |  |
| Male | 128 (53.3) | 48 (24.8) |
| Female | 112 (46.7) | 146 (75.2) |
| **Nationality** (N=240/N=194) |  |  |
| Swiss | 188 (78.3) | 144 (74.2) |
| Portuguese | 10 (4.2) | 8 (4.1) |
| Italian | 14 (5.8) | 14 (7.2) |
| French | 13 (5.4) | 9 (4.6) |
| Others | 15 (6.3) | 19 (9.9) |
| **Ethnicity** (N=240/N=194) |  |  |
| European-Caucasian | 194 (80.1) | 165 (85.1) |
| Other | 46 (19.9) | 29 (14.9) |
| **Marital status** (N=194) |  |  |
| Married or in couple | - | 152 (78.4) |
| Separated, single, widowed | - | 42 (21.6) |
| **Household size** in individuals, mean (SD) (N=194) | - | 3.2 (1.1) |
| **Education** (N=194) |  |  |
| Lower | - | 12 (6.2) |
| Medium | - | 62 (32.0) |
| Higher | - | 120 (61.8) |
| **Household financial situation** (N=194) |  |  |
| High | - | 139 (71.6) |
| Average to poor | - | 40 (20.6) |
| Does not want to answer | - | 15 (7.8) |
| **Crowded household** (N=193) | - | 20 (10.3) |
| **Positive anti-SARS-CoV-2 serology** (N=240/N=194) | 53 (22.1) | 45 (23.2) |
| **Low self-reported HRQoL<sup>1</sup>** (N=235) | 26 (11.0) | - |
| **Self-reported sadness** (N=236) | 84 (35.5) | - |
| **Self-reported loneliness** (N=237) | 55 (23.2) | - |
| **Low parent-reported HRQoL<sup>2</sup>** (N=234) | 28 (11.9) | - |
| **Low parent-reported emotional well-being<sup>2</sup>** (N=234) | 38 (16.2) | - |
| **Average to poor mood** (N=194) | - | 30 (15.5) |

Results are N (%), unless stated otherwise. All variables are self-reported, except serology (see Methods). Other ethnicities include Arab, Asian, African, Indian and South-American. HRQoL stands for health-related quality of life.
<sup>1</sup> Based on the KIDSCREEN-10 scale
<sup>2</sup> Based on the KINDL<sup>®</sup> scale

### Adolescent-reported health-related quality of life, sadness, and loneliness

Overall, 26 (11%) adolescents reported a low HRQoL (Table 1). Multivariable analysis showed that younger adolescents (aOR=0.84; 95%CI: 0.73-0.97) were more likely to present a low HRQoL (Table 2). Referent parents’ positive anti-SARS-CoV2 serology (aOR=3.44; 95%CI: 1.3-9.09) was associated with a lower HRQoL of the adolescent, whereas there was no association with the adolescents’ own serological status (*P* > 0.1). Moreover, engaging in less screen time was associated with a low HRQoL (aOR=0.91; 95%CI: 0.84-0.98), whereas an increase in time spent on social networks since the beginning of the pandemic seemed negatively associated with HRQoL, with borderline significance (aOR=1.82; 95%CI: 0.92-3.66).

**Table 2:** Generalized estimating equations on adolescents’ self-reported Health-Related Quality of Life, HRQoL (KIDSCREEN-10) as well as reported by their parents (KINDL^®^).

| Adolescent-reported HRQoL (KIDSCREEN-10) |  |  |  |  | Parent-reported HRQoL (KINDL®) |  |  |  |
| --- | --- | --- | --- | --- | --- | --- | --- | --- |
|  | N | High<br>N (%) | Low<br>N (%) | Low HRQoL<br>OR (95%CI)<sup>a</sup> |  | High<br>N (%) | Low<br>N (%) | Low HRQoL<br>OR (95%CI)<sup>b</sup> |
| **Age of the adolescent (years)<sup>1</sup>** | 235 | 15.4 (1.6) | 13.8 (4.2) | 0.84 (0.73-0.97)* |  | 15.4 (1.2) | 15.4 (1.1) | 0.92 (0.63-1.35) |
| **Age of the parent (years)<sup>1</sup>** | 235 | 48.4 (5.1) | 48.5 (7.3) | 1.02 (0.92-1.13) |  | 48.3 (5.5) | 48.7 (4.4) | 1.02 (0.94-1.10) |
| **Sex of the adolescent** | 235 |  |  |  | 234 |  |  |  |
| Boy |  | 113 (90.4) | 12 (9.6) | 1 |  | 118 (94.4) | 7 (5.6) | 1 |
| Girl |  | 96 (87.3) | 14 (12.7) | 1.08 (0.57-2.02) |  | 88 (80.7) | 21 (19.3) | 4.21 (1.76-10.10)** |
| **Self-perceived mood of the parent** | 235 |  |  |  | 234 |  |  |  |
| Good |  | 178 (89.9) | 20 (10.1) | 1 |  | 178 (89.4) | 21 (10.6) | 1 |
| Average to poor |  | 31 (83.8) | 6 (16.2) | 1.85 (0.57-5.93) |  | 28 (80.0) | 7 (20.0) | 1.57 (0.62-3.98) |
| **Financial situation of the household** | 235 |  |  |  | 234 |  |  |  |
| Good |  | 156 (90.7) | 16 (9.3) | 1 |  | 152 (88.4) | 20 (11.6) | 1 |
| Average to poor |  | 35 (79.5) | 9 (20.5) | 2.05 (0.75-5.59) |  | 36 (83.7) | 7 (16.3) | 1.06 (0.56-4.59) |
| No answer |  | 18 (94.7) | 1 (5.3) | 0.63 (0.07-5.27) |  | 18 (94.7) | 1 (5.3) | 0.67 (0.08-5.49) |
| **Household size (individuals)<sup>1</sup>** | 235 | 3.2 (1.0) | 3.8 (2.0) | 1.41 (0.81-2.44) |  | 3.3 (1.2) | 3.4 (1.1) | 1.18 (0.80-1.74) |
| **Household density** | 234 |  |  |  | 233 |  |  |  |
| Non-crowded |  | 192 (89.7) | 22 (10.3) | 1 |  | 190 (89.2) | 23 (10.8) | 1 |
| Crowded |  | 16 (80.0) | 4 (20.0) | 2.10 (0.59-6.50) |  | 15 (75.0) | 5 (25.0) | 2.09 (0.78-5.59) |
| **Change in social networks habits** | 229 |  |  |  | 228 |  |  |  |
| Same or less |  | 115 (92.0) | 10 (8.0) | 1 |  | 109 (87.9) | 15 (12.1) | 1 |
| More |  | 88 (84.6) | 16 (15.4) | 1.82 (0.92-3.66) |  | 91 (87.5) | 13 (12.5) | 0.77 (0.30-2.00) |
| **Screen time (hours)<sup>1</sup>** | 229 | 1.8 (2.0) | 1.6 (1.1) | 0.91 (0.84-0.98)* | 228 | 1.9 (2.0) | 1.7 (1.3) | 0.95 (0.85-1.06) |
| **Parent anti-SARS-CoV-2 serology** | 235 |  |  |  | 234 |  |  |  |
| Negative |  | 163 (92.1) | 14 (7.9) | 1 |  | 159 (90.3) | 17 (9.7) | 1 |
| Positive |  | 46 (79.3) | 12 (20.7) | 3.44 (1.30-9.09)* |  | 47 (81.0) | 11 (19.0) | 1.94 (0.64-5.81) |
| **Adolescent anti-SARS-CoV-2 serology** | 235 |  |  |  | 234 |  |  |  |
| Negative |  | 167 (90.8) | 17 (9.2) | 1 |  | 162 (89.0) | 20 (11.0) | 1 |
| Positive |  | 42 (82.4) | 9 (17.6) | 1.12 (0.55-2.27) |  | 44 (84.6) | 8 (15.4) | 1.57 (0.48-5.06) |

Results are odds ratio (OR) and 95% confidence intervals (CI) from multivariable generalized estimating equations adjusted for all covariates in first column.
<sup>a</sup> based on 224 observations, <sup>b</sup> based on 225 observations
\* indicates $P < 0.05$ ; \*\* indicates $P < 0.01$
<sup>1</sup>Descriptive analysis presented as mean (SD); OR applicable for each additional unit of continuous variables

Focusing on psychological distress, 35% of the adolescents reported feeling sad during the previous week, and 23% felt alone (Table 3). Sadness was more likely among girls, compared to boys (aOR=3.29; 95%CI: 1.64-6.57), and was associated with an increase in the use of social networks (aOR=2.05; 95%CI: 1.08-3.88) and an average to poor household financial situation (aOR=2.30; IC95%: 1.00-5.29) with a borderline significance level. Sadness was also associated with the referent parent’s positive anti-SARS-CoV2 serology (aOR=2.38; 95%CI: 1.10-5.14), older age (aOR=1.07; 95%CI: 1.00-1.15) or average to poor mood (aOR=2.81; 95%CI: 1.21-6.56). Female adolescents (aOR=4.15; 95%CI: 2.02-8.51) and those who increased their time spent on social networks (aOR=1.93; 95%CI: 0.96-0.3.90) were more likely to report feeling lonely, while a positive anti-SARS-CoV-2 serology was associated with less loneliness (aOR=0.20; 95%CI: 0.07-0.63) (Table 2).

**Table 3:** Generalized estimating equations on adolescents’ self-reported loneliness and sadness and emotional well-being as reported by their parents

|  | Self-reported sadness |  |  |  | Self-reported Loneliness |  |  |  | Parent-reported low emotional well-being (KINDL®) |  |  |  |
| --- | --- | --- | --- | --- | --- | --- | --- | --- | --- | --- | --- | --- |
|  | N | No N(%) | Yes N(%) | Sadness OR (95% CI)<sup>a</sup> | N | No N(%) | Yes N(%) | Loneliness OR (95% CI)<sup>a</sup> | N | High N(%) | Low N(%) | Low emotional well-being OR (95% CI)<sup>b</sup> |
| **Age of the adolescent (years)<sup>1</sup>** | 236 | 15.4 (1.1) | 14.9 (3.1) | 0.88 (0.76-1.03) | 237 | 15.3 (1.6) | 14.9 (3.2) | 0.95 (0.81-1.11) | 234 | 15.4 (1.2) | 15.3 (1.1) | 0.78 (0.57-1.07) |
| **Age of the parent (years)<sup>1</sup>** | 236 | 47.8 (4.9) | 49.5 (6.0) | 1.07 (1.00-1.15) * | 237 | 48.2 (5.2) | 49.3 (6.0) | 1.05 (0.96-1.15) | 234 | 48.0 (5.2) | 50.2 (5.9) | 1.08 (0.99-1.17) |
| **Sex of the adolescent** | 236 |  |  |  | 237 |  |  |  |  |  |  |  |
| Boy |  | 94 (75.2) | 31 (24.8) | 1 |  | 109 (86.5) | 17 (13.5) | 1 | 234 | 112 (89.6) | 13 (10.4) | 1 |
| Girl |  | 58 (52.3) | 53 (47.7) | 3.29 (1.64-6.57) ** |  | 73 (65.8) | 38 (34.2) | 4.15 (2.02-8.51) ** |  | 84 (77.1) | 25 (22.9) | 3.03 (1.32-6.90) * |
| **Self-perceived mood of the parent** | 236 |  |  |  |  |  |  |  |  |  |  |  |
| Good |  | 138 (69.3) | 61 (30.7) | 1 | 237 | 158 (79.0) | 42 (21.0) | 1 | 234 | 168 (84.4) | 31 (15.6) | 1 |
| Average to poor |  | 14 (37.8) | 23 (62.2) | 2.81 (1.21-6.56) * |  | 24 (64.9) | 13 (35.1) | 1.67 (0.71-3.98) |  | 28 (80.0) | 7 (20.0) | 0.86 (0.29-2.55) |
| **Financial situation of the household** | 236 |  |  |  |  |  |  |  |  |  |  |  |
| Good |  | 115 (66.9) | 57 (33.1) | 1 | 237 | 136 (78.6) | 37 (21.4) | 1 | 234 | 147 (85.5) | 25 (14.5) | 1 |
| Average to poor |  | 22 (48.9) | 23 (51.1) | 2.30 (1.00 -5.29) * |  | 30 (66.7) | 15 (33.3) | 2.37 (0.95-5.85) |  | 32 (74.4) | 11 (25.6) | 2.08 (1.00-7.79) * |
| No answer |  | 15 (78.9) | 4 (21.1) | 0.77 (0.20-2.92) |  | 16 (84.2) | 3 (15.8) | 1.41 (0.36-5.50) |  | 17 (89.5) | 2 (10.5) | 1.12 (0.26-4.81) |
| **Household size (individuals)<sup>1</sup>** | 236 | 3.3 (1.1) | 3.3 (1.3) | 1.16 (0.92-1.45) | 237 | 3.3 (1.1) | 3.4 (1.5) | 1.15 (0.82-1.61) |  | 3.4 (1.2) | 3.1 (1.4) | 0.75 (0.44-1.27) |
| **Household density** | 235 |  |  |  | 236 |  |  |  | 233 |  |  |  |
| Non-crowded |  | 142 (66.0) | 73 (34.0) | 1 |  | 167 (77.3) | 49 (22.7) | 1 |  | 182 (85.4) | 31 (14.6) | 1 |
| Crowded |  | 10 (50.0) | 10 (50.0) | 1.48 (0.48-4.54) |  | 15 (75.0) | 5 (25.0) | 1.13 (0.84-1.52) |  | 13 (65.0) | 7 (35.0) | 3.20 (1.26-8.10) * |
| **Change in social networks habits** | 230 |  |  |  | 231 |  |  |  | 228 |  |  |  |
| Same or less |  | 93(73.8) | 33 (26.2) | 1 |  | 105 (83.3) | 21 (16.7) | 1 |  | 107 (86.3) | 17 (13.7) | 1 |
| More |  | 55 (52.9) | 49 (47.1) | 2.05 (1.08-3.88) * |  | 72 (68.6) | 33 (31.4) | 1.93 (0.96-3.90) |  | 84 (80.8) | 20 (19.2) | 1.33 (0.52-3.36) |
| **Screen time (hours)<sup>1</sup>** | 230 | 1.7 (1.8) | 2.0 (2.0) | 1.04 (0.97-1.11) | 231 | 1.8 (1.8) | 2.1 (2.1) | 1.03 (0.96-1.12) | 228 | 1.9 (2.0) | 1.5 (1.5) | 1.04 (0.96-1.12) |
| **Parent anti-SARS-CoV-2 serology** | 236 |  |  |  | 237 |  |  |  | 234 |  |  |  |
| Negative |  | 121 (68.4) | 56 (31.6) | 1 |  | 139 (78.1) | 39 (21.9) | 1 |  | 148 (84.1) | 28 (15.9) | 1 |
| Positive |  | 31 (52.5) | 28 (47.5) | 2.38 (1.10-5.14) * |  | 43 (72.9) | 16 (27.1) | 2.04 (0.89-5.17) |  | 48 (82.8) | 10 (17.2) | 0.67 (0.22-2.02) |
| **Adolescent anti-SARS-CoV-2 serology** | 236 |  |  |  | 237 |  |  |  |  |  |  |  |
| Negative |  | 116 (63.4) | 67 (36.6) | 1 |  | 135 (73.4) | 49 (26.6) | 1 | 234 | 153 (84.1) | 29 (15.9) | 1 |
| Positive |  | 36 (67.9) | 17 (32.1) | 0.55 (0.24-1.25) |  | 47 (88.7) | 6 (11.3) | 0.20 (0.07-0.63) ** |  | 43 (82.7) | 9 (17.3) | 1.59 (0.60-4.22) |

Results are odds ratio (OR) and 95% confidence intervals (CI) from multivariable generalized estimating equations adjusted for all covariates in first column
<sup>a</sup> based on 229 observations; <sup>b</sup> based on 225 observations
\* indicates $P < 0.05$ ; \*\* indicates $P < 0.01$
<sup>1</sup> Descriptive analysis presented as mean (SD); OR applicable for each additional unit of continuous variables

### Parent-reported Health-related quality of life and emotional well-being

Based on parents’ perception, 28 (12%) adolescents presented a low HRQoL (Table 1). The latter was only associated with adolescents’ sex, girls being more likely to be perceived as having a low HRQoL compared to boys (aOR=4.21; 95%CI: 1.76-10.10; Table 2). When focusing on psychological distress, 38 (16%) adolescents presented a low emotional well-being according to their parents (Table 1), which was associated with being female (aOR=3.03; 95%CI: 1.32-6.90), living in a crowded household (aOR=3.20; 95%CI: 1.26**-**8.10), and having an average to poor financial situation (aOR=2.08; 95%CI: 1.00-7.79; Table 3).

### Comparison of adolescents’ and parents’ perceptions

Based on the dichotomous classification of the KIDSCREEN-10 and the KINDL^®^, adolescents’ and parents’ perceptions matched in 184 (79%) of cases. However, 22 (9.5%) of the adolescents presented a low HRQoL that did not seem identified by their parents. Misperception was more likely among girls compared to boys (aOR**=**2.88; 95%CI: 1.54-5.41) and in households with lower family well-being (aOR**=**0.91; 95%CI: 0.86-0.96; Table 4).

**Table 4:** Risk factors of discrepancies between adolescents’ and parents’ perception of adolescents’ low HRQoL

| Adolescent and parent perception of adolescent HRQoL |  |  |  |  |
| --- | --- | --- | --- | --- |
|  | N | Match<br>N(%) | Mismatch<br>N(%) | Perception mismatch<br>OR (95%CI) |
| **Age of the adolescent (years)<sup>1</sup>** | 232 | 15.4 (1.2) | 15.2 (1.1) | 0.86 (0.68-1.16) |
| **Age of the parent (years)<sup>1</sup>** | 232 | 48.4 (5.3) | 48.4 (5.8) | 1.02 (0.96-1.10) |
| **Sex of the adolescent** | 232 |  |  |  |
| Boy |  | 108 (87.1) | 16 (12.9) | 1 |
| Girl |  | 76 (70.4) | 32 (29.6) | 2.88 (1.54-5.41) ** |
| **Self-perceived mood of the parent** | 232 |  |  |  |
| Good |  | 161 (81.7) | 36 (18.3) | 1 |
| Average to poor |  | 23 (65.7) | 12 (34.3) | 1.91 (0.67-5.39) |
| **Financial situation of the household** | 232 |  |  |  |
| Good |  | 135 (78.9) | 36 (21.1) | 1 |
| Average to poor |  | 32 (76.2) | 10 (23.8) | 1.03 (0.68-2.88) |
| No answer |  | 17 (89.5) | 2 (10.5) | 0.37 (0.07-1.90) |
| **Household size (individuals)<sup>1</sup>** | 232 | 3.2 (1.0) | 3.6 (1.7) | 1.24 (0.85-1.82) |
| **Household density** | 231 |  |  |  |
| Non-Crowded |  | 172 (81.5) | 39 (18.5) | 1 |
| Crowded |  | 11 (55.0) | 9 (45.0) | 2.51 (0.76-8.09) |
| **Change in social networks habits** | 226 |  |  |  |
| Same or less |  | 99 (80.5) | 24 (19.5) | 1 |
| More |  | 79 (76.7) | 24 (23.3) | 0.88 (0.42-1.88) |
| **Screen time (hours)<sup>1</sup>** | 226 | 1.9 (2.0) | 1.6 (1.2) | 0.93 (0.87-1.01) |
| **Serology result of the parent** | 232 |  |  |  |
| Negative |  | 145 (82.9) | 30 (17.1) |  |
| Positive |  | 39 (68.4) | 18 (31.6) | 1.72 (0.67-4.37) |
| **Serology result of the adolescent** | 232 |  |  |  |
| Negative |  | 148 (81.3) | 34 (18.7) | 1 |
| Positive |  | 36 (72.0) | 14 (28.0) | 2.54 (0.98-6.62) |
| **Family well-being score<sup>1</sup>** | 232 | 66.3 (7.8) | 58.8 (10.0) | 0.91 (0.86-0.96) ** |

Results are odds ratio (OR) and 95% confidence intervals (CI) from multivariable generalized estimating equations adjusted for all covariates in first column based on 225 observations. Perception mismatch is coded as 1 if low HRQoL from adolescents' and parents' perception do not match and 0 otherwise.
\* indicates $P < 0.05$ ; \*\* indicates $P < 0.01$ .
<sup>1</sup> Descriptive analysis presented as mean (SD); OR applicable for each additional unit of continuous variables

## 4. DISCUSSION

In this population-based study, we observed that a meaningful proportion of adolescents reported a low well-being HRQoL and some psychological distress during the second COVID-19 wave in Switzerland, 8 to 9 months after the start of the pandemic. Adolescents’ self-reported well-being was generally corroborated by their parent’s observation. Risk factors for a low well-being from the adolescents’ perspective included a younger age and a positive anti-SARS-CoV-2 serology of the referent parent, while being a girl was the only risk factor for low HRQoL as perceived by parents. Girls compared to boys, and adolescents living in households with lower family well-being, were at higher risk of misperception by their parents. When looking at psychological distress, risk factors for adolescent-reported sadness or loneliness included being a girl, spending more time on social networks or living in a household with a disadvantaged financial situation, as well as characteristics of the referent parent such as older age, positive anti-SARS-CoV-2 serology and lower mood.

Compared to the Swiss pre-pandemic reference level where the KIDSCREEN-10 median corresponded to a score of 80 [16,22], adolescents presented lower HRQoL in this study with a median of 72.5 (MAD = 14.8), which may be partly explained by the impact of the pandemic. This is in line with results from other studies in China [23], Brazil [24], Germany [17], Australia [25], India [26], Spain [27] and Italy [27] which reported high levels of adolescent stress, worry and anxiety during the pandemic, likely due to restrictive measures to contain COVID-19 spread and the worrying pandemic environment. Estimates of sadness and loneliness among adolescents were lower in the present study than what reported in other studies in China [23], India [26], Spain [27] and Italy [27]. However, most previous studies were based on convenience samples and were not population-based. Further, in Switzerland measures to contain virus spread were comparatively less strict than in other countries. For example, inland travel, small gatherings and outdoor sports were never forbidden [14,28]. Finally, during the study period sanitary measures were regularly changing in Geneva, including restrictions on extra-curricular activities and mask wearing at schools, but were never as strict as during the first pandemic wave when schools were closed and activities suspended [14]. The psychological distress estimates are thus likely to be lower than what would have been observed during the first wave, when most other studies have been conducted [23,26,27].

Lower well-being was associated with a younger age, as also found by Ravens-Sieberer et al [17] and with being a girl, as generally observed [25,29]. Parental lower mood was a risk factor for adolescent psychological distress, which is consistent with studies showing that parent’s mental health directly impacts children’s functioning [12]. Living in a crowded household was associated with adolescents’ psychological distress, which could be explained by difficulties to maintain privacy and a personal space at home [30]. This aspect has possibly worsened since the start of the pandemic as household members were likely to spend more time at home than usual. On the opposite, household size was not associated with adolescents’ well-being, consistent with other findings [31].

In accordance with warnings issued by psychologists, an increase in time spent on social networks was associated with sadness and loneliness [32]. Causality could be bidirectional. As a consequence of other activities being restricted, such as sport or music lessons, adolescents may have felt lonely, sad and bored, and thus have spent more time on social media. Conversely, it may have increased their exposure to alarming and contradictory information and affected their well-being [33]. Interestingly, overall screen time was not related to psychological distress, suggesting that examining the type of online activities may be of more significance than the overall screen time [34].

Another unexpected finding was that adolescents’ low HRQoL was associated with a positive anti-SARS-CoV-2 serology of the parent but not with their own serological result. In line with other results [35], this may reflect that adolescents were more worried or impacted by their parents health rather than by their own during the COVID-19 pandemic. It may also mirror the impact on adolescents of potential difficult circumstances linked with a parent infected by SARS-CoV-2, such as a severe disease or long-term effects due to long COVID [36]. Adolescents’ negative anti-SARS-CoV-2 serology was associated with loneliness. A possible explanation could be that adolescents respecting social distancing measures more carefully were less infected but also felt lonelier, although this remains speculative at this stage.

An association between poor socioeconomic conditions and adolescents’ psychological distress was probably already present before the COVID-19 pandemic [37]. However, its significance was likely to increase as restrictions may have resulted in fewer quiet spaces, less resources to help with school work or increased likelihood of financial instability for adolescents in general, and more particularly vulnerable ones [13]. Longitudinal studies are necessary to assess the effect of the pandemic on adolescents’ mental health over time. More specifically, the impact of socioeconomic conditions should be examined, as deprivation could result in lower resilience and constitute a risk factor for persistent low well-being.

Adolescents’ and parents’ perceptions of adolescents’ HRQoL matched in most cases. This finding is important as parents play a role in the early detection of psychological distress among their children and in care seeking [38]. Especially during the COVID-19 pandemic, parents play a key role in reducing the negative psychological impact on their children, for example through attention and supportive behaviours, such as keeping regular routines as much as possible or providing clear explanations about the situation [39]. However, 9.5% of the adolescents presented a low HRQoL that did not seem to be identified by their parents. Misperception was more likely among girls and adolescents living in families with lower well-being; these adolescents may represent a vulnerable group whose mental health issues are under-recognized.

The study presents several limitations. First, it is a cross-sectional study relying on self-reported data, without any pre-pandemic hindsight coming from the same sample. It does not allow us to firmly conclude whether the reported adolescents’ low quality of life is caused by the pandemic. The sample size is quite small, which limited statistical power. Finally, though randomly invited, study participants come from a higher socioeconomic background than the Geneva population. As a consequence, estimates of low well-being and psychological distress may have been underestimated. This study also presents several strengths. First, it relies on a randomly selected population-based sample. Thanks to the family-based design, data comes from both adolescents and their parents, which enables comparisons of both perceptions. Furthermore, previous SARS-CoV-2 infection is assessed with an objective measure. Finally, the demographic and socioeconomic characteristics of the sample are assessed through a wide range of indicators, at the individual and household levels.

## 5. CONCLUSION

Amid the COVID-19 second wave in Geneva, a meaningful share of adolescents reported low psychological well-being, especially those living in unfavourable family environments, including crowded households, in poor financial situation or with distressed parents. Tailored measures, especially targeting these vulnerable adolescents, are necessary and could include free psychological consultations or resources for parents to react adequately to this special situation. Finally, monitoring is necessary to evaluate the long-term effects of the pandemic on adolescents’ mental health and inform decision-makers as the pandemic continues. Indeed some of the impacts may be temporary thanks to the lifting of restrictions, while others may persist over time, especially among vulnerable individuals.

## Data Availability

Our data are accessible to researchers upon reasonable request for data sharing to the corresponding author.

## Acknowledgment

We are grateful to the staff of the Unit of Population Epidemiology of the HUG Division of Primary Care Medicine as well as to all the participants whose contributions were invaluable to the study.

